# Targeted Sequencing of the 9p21.3 Region Reveals Association with Reduced Disease risks in Ashkenazi Jewish Centenarians

**DOI:** 10.1101/2023.05.19.23290237

**Authors:** Yizhou Zhu, Seungjin Ryu, Archana Tare, Nir Barzilai, Gil Atzmon, Yousin Suh

**Affiliations:** Department of Obstetrics and Gynecology, Columbia University, New York, NY10032, USA; Department of Genetics, Albert Einstein College of Medicine, Bronx, NY10461, USA; Department of Medicine, Albert Einstein College of Medicine, Bronx, NY10461, USA; Department of Genetics and Development, Columbia University, New York, NY10461, USA

**Keywords:** Longevity, Aging, Age-related disease, 9p21, Centenarians, Population Genomics

## Abstract

Genome-wide association studies (GWAS) have pinpointed the chromosomal locus 9p21.3 as a genetic hotspot for various age-related disorders. Common genetic variants in this locus are linked to multiple traits, including coronary artery diseases, cancers, and diabetes. Centenarians are known for their reduced risk and delayed onset of these conditions. To investigate whether this evasion of disease risks involves diminished genetic risks in the 9p21.3 locus, we sequenced this region in an Ashkenazi Jewish centenarian cohort (centenarians: n=450, healthy controls: n=500). Risk alleles associated with cancers, glaucoma, CAD, and T2D showed a significant depletion in centenarians. Furthermore, the risk- and non-risk genotypes are linked to two distinct low-frequency variant profiles, enriched in controls and centenarians, respectively. Our findings provide evidence that the extreme longevity cohort is associated with collectively lower risks of multiple age-related diseases in the 9p21.3 locus.

## Main Text

Longevity is a multifaceted phenotype influenced by a combination of environmental and genetic factors. Twin studies have demonstrated that longevity is moderately heritable (estimated at 20-30%), with genetic factors playing a more significant role in achieving extended longevity at higher ages^1^. Extremely long-lived individuals often exhibit healthy aging characteristics, such as the absence or delayed onset of age-related diseases, suggesting that they may be genetically protected from age-related disease risks^2^. However, previous research has shown that disease risk alleles identified through genome-wide association studies (GWAS) are commonly found in longevity cohorts^3^. This implies the existence of alternative mechanisms for controlling these disease risks, such as the presence of protective rare variants.

The 9p21.3 non-coding locus, located upstream of the INK4/ARF (CDKN2A/B) genes, remains one of the most consistently replicated GWAS hotspots^4,5^. This locus has been linked to risk of multiple age-related diseases, including cardiovascular diseases (CVD), type 2 diabetes (T2D), glaucoma, and multiple cancers^6-12^. In contrast to the strong association of 9p21.3 with age-related diseases, fewer studies have explored the locus’s relationship with longevity. A genome-wide association study of the New England Centenarian cohort reported a weak association signal for rs1063192, a 3’ UTR variant on CDKN2B, which has also been linked to glaucoma^13^. The UK Biobank identified an association between rs1556516, a CAD-related variant, and parental longevity^14^. Another top variant associated with coronary artery disease, rs1333049, was found to be connected to longevity in a Spanish centenarian cohort as well as the Wellderly healthy aging cohort^15,16^. However, this association was not confirmed in two Japanese studies performed by independent groups^15,17^. It is important to note that the majority of studies have focused on genotyping common disease variants. Consequently, the implications of rare variants in this region with respect to longevity remain largely uncharacterized.

To comprehensively investigate the association between extreme longevity and the 9p21.3 genotype, we conducted a sequencing study of this locus in Ashkenazi Jewish (AJ) centenarians (n=450; mean age = 98 for cases and n=500; mean age = 73 for controls). The AJ population is genetically homogenized^18,19^. Utilizing pooled capture sequencing, we sequenced the 230 kb GWAS interval (chr9: 21,950,000-22,180,000, hg19) with an average 30x depth^20^. The accuracy of the sequencing results was validated by genotyping (r^2^ >=0.99, **Figure S1, Table S1**).

We identified 2,216 variants, including 2,056 single nucleotide polymorphisms (SNPs) and 160 indels (**Table 1A**). Comparing these variants with the current SNP database (SNP149) revealed that 785 out of 2,216 (35.4%) were novel, comprising 664 SNPs and 47 indels. Among all novel variants, 95% (743) were rare (minor allele frequency <1%), and 78% (616) were singletons.

**Table1.**
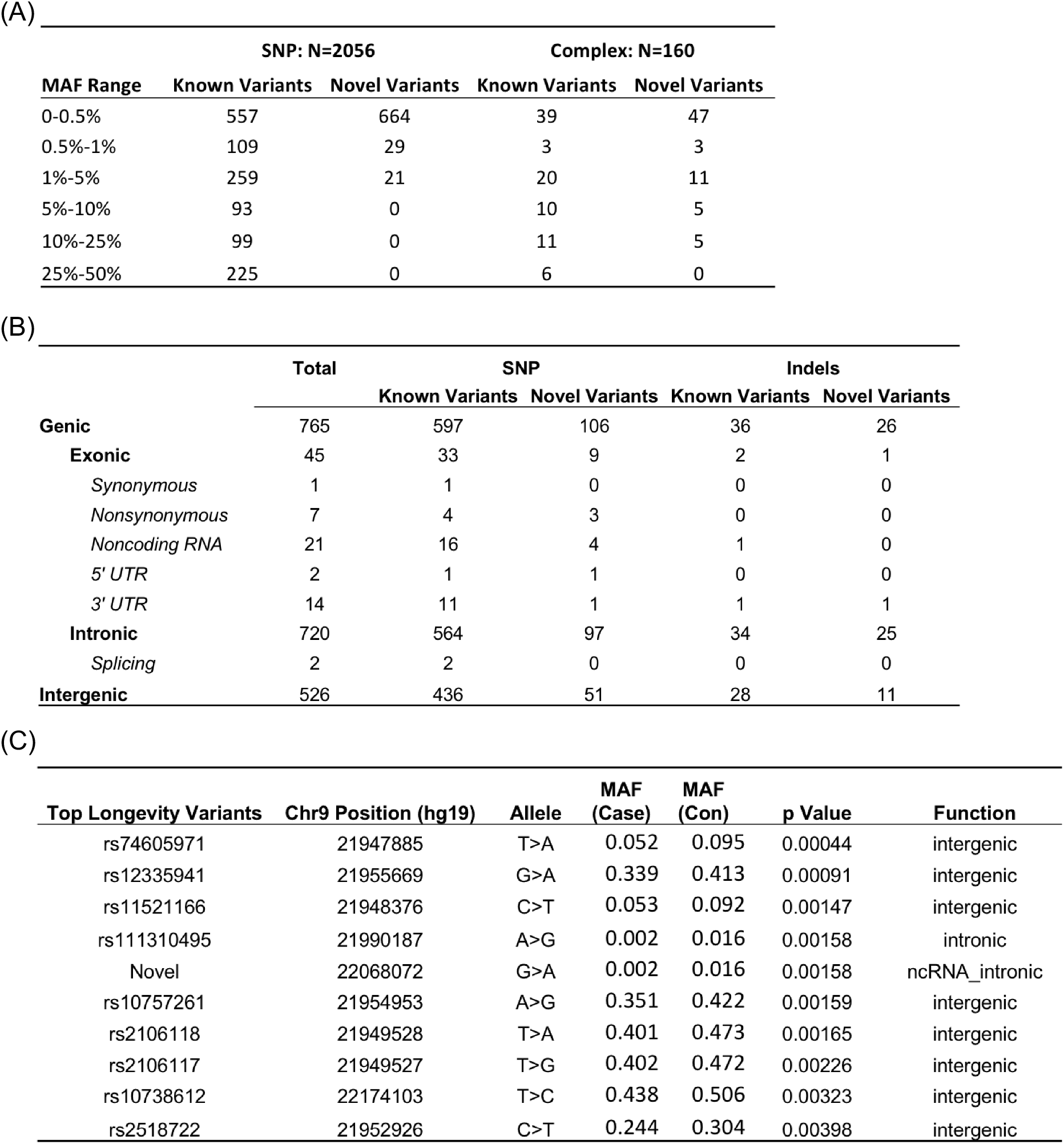
(A) Summary of variants identified by sequencing of 9p21.3 in 450 centenarians and 500 controls arranged by minor allele frequency ranges. Variant annotation was performed with dbSNP database 149. (B) Functional annotation of 1064 identified variants with at least 4 allele counts. Genes considered include CDKN2A, CDKN2B, and non-coding transcript ANRIL (CDKN2BAS1). (C) List of top 10 longevity-associated variants.

| MAF Range | SNP: N=2056 |  | Complex: N=160 |  |
| --- | --- | --- | --- | --- |
|  | Known Variants | Novel Variants | Known Variants | Novel Variants |
| 0-0.5% | 557 | 664 | 39 | 47 |
| 0.5%-1% | 109 | 29 | 3 | 3 |
| 1%-5% | 259 | 21 | 20 | 11 |
| 5%-10% | 93 | 0 | 10 | 5 |
| 10%-25% | 99 | 0 | 11 | 5 |
| 25%-50% | 225 | 0 | 6 | 0 |

(B)
|  | Total | SNP |  | Indels |  |
| --- | --- | --- | --- | --- | --- |
|  |  | Known Variants | Novel Variants | Known Variants | Novel Variants |
| **Genic** | 765 | 597 | 106 | 36 | 26 |
| **Exonic** | 45 | 33 | 9 | 2 | 1 |
| *Synonymous* | 1 | 1 | 0 | 0 | 0 |
| *Nonsynonymous* | 7 | 4 | 3 | 0 | 0 |
| *Noncoding RNA* | 21 | 16 | 4 | 1 | 0 |
| 5' UTR | 2 | 1 | 1 | 0 | 0 |
| 3' UTR | 14 | 11 | 1 | 1 | 1 |
| **Intronic** | 720 | 564 | 97 | 34 | 25 |
| *Splicing* | 2 | 2 | 0 | 0 | 0 |
| **Intergenic** | 526 | 436 | 51 | 28 | 11 |

**Table1** (A) Summary of variants identified by sequencing of 9p21.3 in 450 centenarians and 500 controls arranged by minor allele frequency ranges.
| Top Longevity Variants | Chr9 Position (hg19) | Allele | MAF (Case) | MAF (Con) | p Value | Function |
| --- | --- | --- | --- | --- | --- | --- |
| rs74605971 | 21947885 | T>A | 0.052 | 0.095 | 0.00044 | intergenic |
| rs12335941 | 21955669 | G>A | 0.339 | 0.413 | 0.00091 | intergenic |
| rs11521166 | 21948376 | C>T | 0.053 | 0.092 | 0.00147 | intergenic |
| rs111310495 | 21990187 | A>G | 0.002 | 0.016 | 0.00158 | intronic |
| Novel | 22068072 | G>A | 0.002 | 0.016 | 0.00158 | ncRNA_intronic |
| rs10757261 | 21954953 | A>G | 0.351 | 0.422 | 0.00159 | intergenic |
| rs2106118 | 21949528 | T>A | 0.401 | 0.473 | 0.00165 | intergenic |
| rs2106117 | 21949527 | T>G | 0.402 | 0.472 | 0.00226 | intergenic |
| rs10738612 | 22174103 | T>C | 0.438 | 0.506 | 0.00323 | intergenic |
| rs2518722 | 21952926 | C>T | 0.244 | 0.304 | 0.00398 | intergenic |

We functionally annotated 1,291 variants with at least four counts of minor alleles (**Table 1B**). The vast majority of these variants were intronic (720, 55.8%) or intergenic (526, 40.7%). Among the 45 (3.5%) exonic variants, 21 were located in ANRIL and 10 in 5’ or 3’ UTR. We identified seven non-synonymous and one synonymous SNPs, including three novel variants: one in the CDKN2A gene (chr9:21974675 A>C, V51G, MAF=0.84%), and two in CDKN2B (chr9:22006101 C>T, R101Q, MAF=0.21% and chr9 22008790 C>A, G55W, MAF=0.26%). These candidate functional variants were not found to be associated with longevity (p>0.05).

We identified 84 variants associated with longevity based on nominal p-values (**Table 1C**). The majority of top hit SNPs were situated downstream of CDKN2A (**Figure 1A**). Among the GWAS-reported SNPs, the variant with the highest significance was rs4977756 (p=0.019, OR=0.78), which has been linked to glaucoma^21^. This variant was also reported in a previous longevity iGWAS study using the same cohort^22^. Most GWAS variants were not found to be significantly associated with longevity in this study, including rs1333049 (p=0.36, OR=0.92), the strongest coronary artery disease variant (**Table S2**). However, a lower odds ratio for the risk allele was consistently observed for centenarians across all but one trait, suggesting a trend of combined risk variant depletion (**Figure 1B**). The only exception was glioma (rs1412829), which could be attributed to its non-risk allele being linked to the risks of other diseases, such as glaucoma and cardiovascular traits.

**Figure 1.**
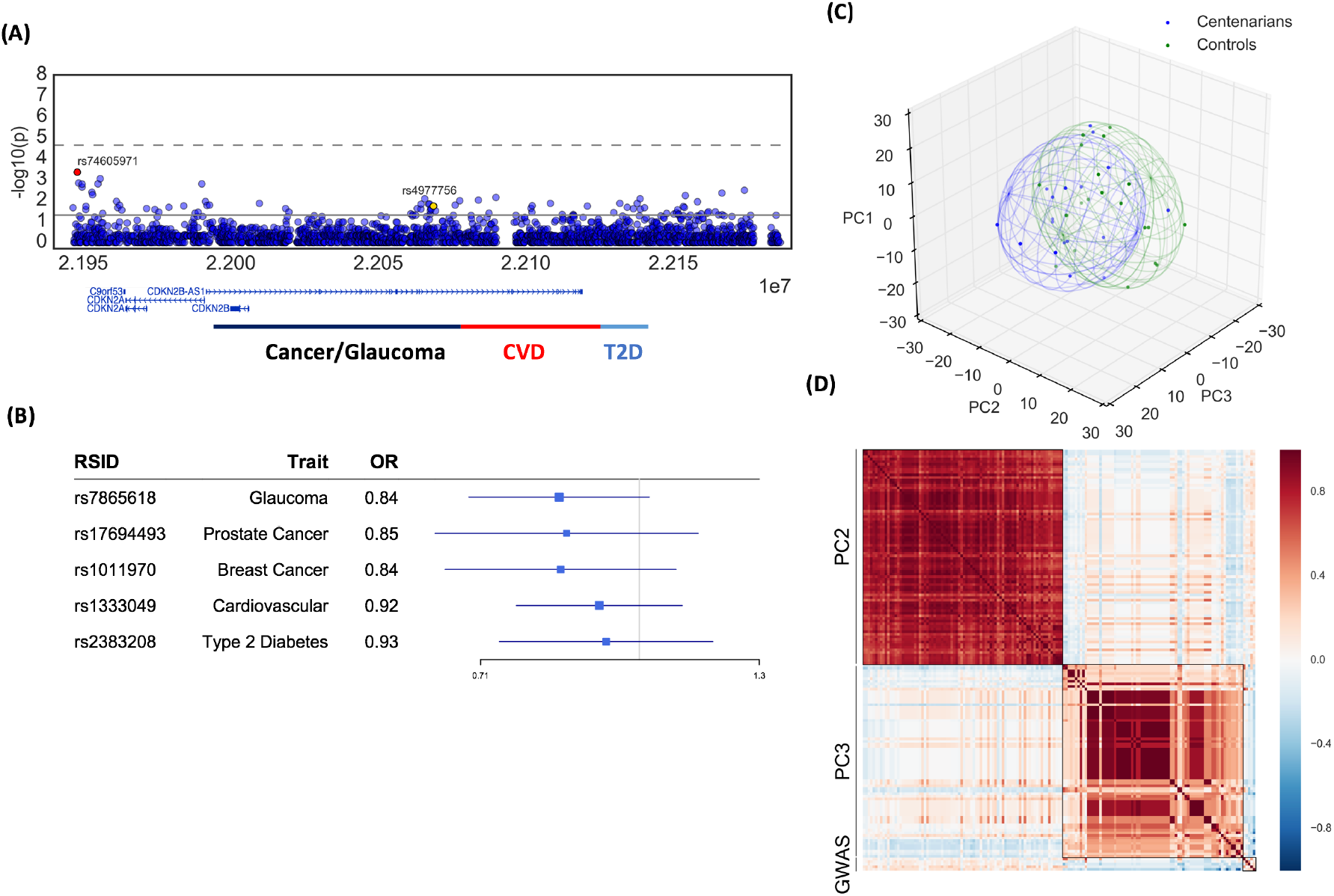
(A) Regional plots of 9p21.3 genotype-longevity associations. Solid line and dash line represent nominal p and adjusted p threshold, respectively. Subdivisions of the locus with enriched GWAS variants are indicated at bottom. (B) Forest plot for five distinct GWAS SNPs representing major disease traits associated with the 9p21 locus (C) Distribution of first three principle components from PCA for the 18 centenarian pools and 20 control pools. Ellipsoids indicate distribution confidence interval. (D) Correlation matrix between the high importance SNPs from PC2 and PC3 and the GWAS risk variants in (B).

To statistically test the significance of GWAS risk allele depletion in centenarians, we reduced the complete list of GWAS variants to five uncorrelated (r^2^ ≤ 0.1) tag SNPs (**Figure S2**)^23^. Each tag SNP represented its linked variants, typically associated with the same traits, and we examined their combined distribution using a permutation test. The result was significant (p < 10^−4^), suggesting that the overall age-related disease risks in the 9p21.3 region were lower in centenarians.

With individual SNP tests demonstrating weak yet overall significant associations between disease risk variants and longevity, we next employed machine learning to determine if the depletion of risk alleles for each trait was independent. Using random forest and boosting techniques^24^, we found that variants downstream of CDKN2A and around rs4977756 both showed high feature importance, suggesting that the associations of these two regions with longevity are independent (**Figure S3**).

Meanwhile, principal component analysis (PCA) revealed major differences of the 9p21.3 genotype in control and longevity cohorts within the first three principal components (**Figure 1C**). We applied a linear support vector classifier (SVC) and found the first 3 principal components (PCs) significantly distinguished the two cohorts (accuracy = 74%, p = 0.049, permutation test). Further analysis of individual components showed that the centenarian group had notably lower PC2 (p = 0.036, Mann-Whitney U test) and higher PC3 (p = 0.089) values.

Upon examining the feature importance in these components, we discovered that PC2 and PC3 were characterized by two distinct groups of linked low-frequency common variants (MAF < 5% for both PC2 and PC3) (**Figure S4, S5A**). Interestingly, the minor alleles of variants with high importance in PC2 were associated with the risk alleles of all five flag GWAS SNPs, while those in PC3 were linked to the non-risk alleles (**Figure 2D**). This finding suggests that PC2 and PC3 represent low-frequency genotypes associated with high and low combinatorial disease risks, respectively. Hence, the low PC2 and high PC3 scores in centenarians indicate an enrichment of genotypes with overall reduced genetic risks for age-related diseases in the 9p21.3 region, which aligns with the analysis of GWAS SNPs.

To assess the association of clustered rare variants with longevity, we performed sequence kernel association test (SKAT)^25^. After breaking down the locus by position of the genes and performed SKAT separately, the association of CDKN2A downstream region was found significant when the direction of variants was considered (**Table S3**). We also performed SKAT on potential regulatory elements in 127 epigenomes from Roadmap Project. No significant association was identified (data not shown). Nevertheless, the minor allele of the majority of variants in this region, both rare and common, was depleted in centenarians, indicating a possible deleterious role of alternative alleles in longevity for variants in this region (**Figure S5B**).

In this study, we conducted a comprehensive sequencing analysis of the 9p21.3 locus, which is associated with multiple age-related phenotypes, in Ashkenazi Jewish centenarians. To our knowledge, this study is the first to extensively characterize the association of all genetic variants in this locus with extreme longevity in a significant cohort size. We identified moderate associations between multiple GWAS risk variants in 9p21.3 and longevity, with the strongest signal originating from rs4977756, a variant reported to be associated with glaucoma risks^21^. Notably, rs4977756 is in high LD with CAD variants (R^2^ = 0.41 in Europeans) and located at the junction between cancer/glaucoma and CAD blocks. This LD block junction region represents the strongest longevity hotspot within the 9p21.3 GWAS locus (**Figure 1A**). Together with the result that the depletion of risk alleles was moderate but consistent for all age-related disorder variants, our data suggest that instead of potently evading the risk of one particular trait associated with 9p21.3, the Ashkenazi Jewish centenarians may carry an overall lower genetic risk at this locus.

Consistent with our findings from single variant analysis, we identified two distinct variant groups that are either enriched or depleted in centenarians. Despite their low minor allele frequencies, both variant groups showed strong correlations in sample distribution (**Figure 2D**). Although further haplotyping is required for confirmation, such patterns strongly suggest that the minor alleles of these variants belong to one or a few haplotypes. We demonstrated that these two variant groups are associated with combined GWAS non-risk and risk alleles, which are enriched and depleted in centenarians, respectively. These findings indicate the presence of rare high and low combined disease risk haplotypes in the 9p21.3 region, which are respectively negatively and positively selected in centenarians.

Despite being one of the earliest identified GWAS loci, the mechanism by which 9p21.3 contributes to disease risk remains largely unclear. It has been demonstrated that the non-coding variants within this locus have regulatory functions and alter the expression levels of neighboring genes, including INK4/ARF and the long non-coding RNA transcript CDKN2B-AS1^26,27^. By sequencing the 9p21.3 locus, we provide a comprehensive list of variants associated with longevity, which serves as a valuable resource for further study of the regulatory mechanisms of this locus.

## Experimental Procedures

### Subjects

The Ashkenazi Centenarian cohort has been described in our previous studies ^18,19^. Briefly, 450 centenarians (mean age: 98) and 500 controls (mean age: 73) with no family record of longevity were involved in the study. Genomic DNA was extracted from whole blood samples of participants and subjected to whole genome amplification using GenomiPhi V2 DNA Amplification kits (GE Healthcare Life Sciences) prior to capture genome sequencing.

### Pooled Target Capture Sequencing

The pooled capture-seq technique was performed as described previously ^20^. In short, genomic DNA samples were divided into groups of 25 and pooled equimolarly. This resulted in 18 pools of centenarians and 20 pools of controls. Using one microgram of pooled DNA, the sequencing library was prepared following Illumina’s Truseq library protocol. The 9p21.3 region (chr9:21950000-22180000, hg19) was specifically captured using the Nimblegen SeqCap EZ Choice library target capture kit according to the manufacturer’s instructions. Sequencing was performed on the Illumina HiSeq2500 platform with an average depth of 30x per individual. Sequencing results were aligned to the human genome hg19 using BWA. After pre-processing with PICARD tools (http://broadinstitute.github.io/picard), multi-sample variant calling was achieved using CRISP ^28^. The variants were annotated to dbSNP149 and refGene for functional annotation/prediction. ANNOVAR was used for functional annotation annotation ^29^.

### Genotyping

Genomic regions with variants subject to genotyping were PCR amplified from genomic DNA using FastStart Taq DNA polymerase (Roche). Genotyping was performed on the Sequenom MassARRAY platform following the manufacturer’s instructions. Genotype calls were performed using TYPER 4.0, the coupled analysis software for the assay, with default call thresholds.

### Statistical Analysis

Allele frequencies for each variant in the centenarian and control groups were calculated, and statistical inference of association was determined using Fisher’s exact tests. For rare variant analysis, the Sequence Kernel Association Test (SKAT) ^25^ and its derivative methods (SKAT-O, SKAT-C) were applied to the entire sequenced interval, as well as subdivisions based on gene locations. Reported p-values were adjusted for multiple testing with Benjamini-Hochberg correction. For permutation tests for combined depletion of risk variants, identification tags for pools were permutated 10,000 times. For each permutated dataset, the Z-scores of the five GWAS SNPs shown in Figure 1B were calculated for the centenarian group. The number of incidences where the sum of Z-scores was equal to or lower than the actual centenarian group were counted and reported as p-values after division by the total number of trials.

The Python Sklearn package was utilized for the feature selection study. Before initiating the learning process, the allele frequency matrix was pre-processed by 1) adjusting the allele count to the minor allele (for cases where the major allele was the alternative), 2) removing singletons and doubletons, and 3) normalizing to Z-score. Random Forest and Gradient Tree Boosting classifiers were implemented using the RandomForestClassifier (n_estimator=10000) and GradientBoostingClassifier (n_estimator=5000, learning_rate=0.02) in the ensemble module, respectively. Principal Component Analysis (PCA) was performed using the PCA function in the decomposition module. A linear Support Vector Classifier (SVC) was built to separate control and centenarian pools using the first 3 PC values with the SVC function in the svm module. The significance of separation was tested by permuting the pool’s identification tags 10,000 times and counting instances where classification accuracy was equal to or higher than the actual data. The python seaborn.heatmap function was used to generate a correlation matrix heatmap. A threshold of 0.05 was applied for PC2 and PC3 to select the important feature SNPs.

## Data Availability

All data produced in the present study are available upon reasonable request to the authors.

## Author Contribution

Conceptualization: YZ, NB, YS; Methodology: YZ, SR, AT; Analysis: YZ; Resource: GA; Drafting: YZ, YS; Editing: all authors

## Data Availability

Data available on request from the authors.

## Ethics Approval

The study was approved by the Committee on Clinical Investigations of the Albert Einstein College of Medicine.

## Reference

1 v, B. H. J. et al. Genetic influence on human lifespan and longevity. Hum Genet 119, 312–321, doi:10.1007/s00439-006-0144-y (2006).

2 Perls, T. T. The different paths to 100. Am J Clin Nutr 83, 484S–487S, doi:10.1093/ajcn/83.2.484S (2006).

3 Brooks-Wilson, A. R. Genetics of healthy aging and longevity. Hum Genet 132, 1323–1338, doi:10.1007/s00439-013-1342-z (2013).

4 Jeck, W. R., Siebold, A. P. & Sharpless, N. E. Review: a meta-analysis of GWAS and age-associated diseases. Aging Cell 11, 727–731, doi:10.1111/j.1474-9726.2012.00871.x (2012).

5 Hannou, S. A., Wouters, K., Paumelle, R. & Staels, B. Functional genomics of the CDKN2A/B locus in cardiovascular and metabolic disease: what have we learned from GWASs? Trends Endocrinol Metab 26, 176–184, doi:10.1016/j.tem.2015.01.008 (2015).

6 Helgadottir, A. et al. The same sequence variant on 9p21 associates with myocardial infarction, abdominal aortic aneurysm and intracranial aneurysm. Nat Genet 40, 217–224, doi:10.1038/ng.72 (2008).

7 Cugino, D. et al. Type 2 diabetes and polymorphisms on chromosome 9p21: a meta-analysis. Nutr Metab Cardiovasc Dis 22, 619–625, doi:10.1016/j.numecd.2010.11.010 (2012).

8 Wiggs, J. L. et al. Common variants at 9p21 and 8q22 are associated with increased susceptibility to optic nerve degeneration in glaucoma. PLoS Genet 8, e1002654, doi:10.1371/journal.pgen.1002654 (2012).

9 Rahmioglu, N. et al. Genetic variants underlying risk of endometriosis: insights from meta-analysis of eight genome-wide association and replication datasets. Hum Reprod Update 20, 702–716, doi:10.1093/humupd/dmu015 (2014).

10 Wrensch, M. et al. Variants in the CDKN2B and RTEL1 regions are associated with high-grade glioma susceptibility. Nat Genet 41, 905–908, doi:10.1038/ng.408 (2009).

11 Sherborne, A. L. et al. Variation in CDKN2A at 9p21.3 influences childhood acute lymphoblastic leukemia risk. Nat Genet 42, 492–494, doi:10.1038/ng.585 (2010).

12 Samani, N. J. et al. Genomewide association analysis of coronary artery disease. N Engl J Med 357, 443–453, doi:10.1056/NEJMoa072366 (2007).

13 Kotake, Y. et al. Long non-coding RNA ANRIL is required for the PRC2 recruitment to and silencing of p15(INK4B) tumor suppressor gene. Oncogene 30, 1956–1962, doi:10.1038/onc.2010.568 (2011).

14 Pilling, L. C. et al. Human longevity: 25 genetic loci associated in 389,166 UK biobank participants. Aging (Albany NY) 9, 2504–2520, doi:10.18632/aging.101334 (2017).

15 Pinos, T. et al. The rs1333049 polymorphism on locus 9p21.3 and extreme longevity in Spanish and Japanese cohorts. Age (Dordr) 36, 933–943, doi:10.1007/s11357-013-9593-0 (2014).

16 Erikson, G. A. et al. Whole-Genome Sequencing of a Healthy Aging Cohort. Cell 165, 1002–1011, doi:10.1016/j.cell.2016.03.022 (2016).

17 Congrains, A. et al. Disease-associated polymorphisms in 9p21 are not associated with extreme longevity. Geriatr Gerontol Int 15, 797–803, doi:10.1111/ggi.12346 (2015).

18 Shlush, L. I. et al. Ashkenazi Jewish centenarians do not demonstrate enrichment in mitochondrial haplogroup J. PLoS One 3, e3425, doi:10.1371/journal.pone.0003425 (2008).

19 Ryu, S., Atzmon, G., Barzilai, N., Raghavachari, N. & Suh, Y. Genetic landscape of APOE in human longevity revealed by high-throughput sequencing. Mech Ageing Dev 155, 7–9, doi:10.1016/j.mad.2016.02.010 (2016).

20 Ryu, S., Han, J., Norden-Krichmar, T. M., Schork, N. J. & Suh, Y. Effective discovery of rare variants by pooled target capture sequencing: A comparative analysis with individually indexed target capture sequencing. Mutat Res 809, 24–31, doi:10.1016/j.mrfmmm.2018.03.007 (2018).

21 Burdon, K. P. et al. Genome-wide association study identifies susceptibility loci for open angle glaucoma at TMCO1 and CDKN2B-AS1. Nat Genet 43, 574–578, doi:10.1038/ng.824 (2011).

22 Fortney, K. et al. Genome-Wide Scan Informed by Age-Related Disease Identifies Loci for Exceptional Human Longevity. PLoS Genet 11, e1005728, doi:10.1371/journal.pgen.1005728 (2015).

23 Machiela, M. J. & Chanock, S. J. LDlink: a web-based application for exploring population-specific haplotype structure and linking correlated alleles of possible functional variants. Bioinformatics 31, 3555–3557, doi:10.1093/bioinformatics/btv402 (2015).

24 Ogutu, J. O., Piepho, H. P. & Schulz-Streeck, T. A comparison of random forests, boosting and support vector machines for genomic selection. BMC Proc 5 Suppl 3, S11, doi:10.1186/1753-6561-5-S3-S11 (2011).

25 Wu, M. C. et al. Rare-variant association testing for sequencing data with the sequence kernel association test. Am J Hum Genet 89, 82–93, doi:10.1016/j.ajhg.2011.05.029 (2011).

26 Harismendy, O. et al. 9p21 DNA variants associated with coronary artery disease impair interferon-gamma signalling response. Nature 470, 264–268, doi:10.1038/nature09753 (2011).

27 Almontashiri, N. A. et al. 9p21.3 Coronary Artery Disease Risk Variants Disrupt TEAD Transcription Factor-Dependent Transforming Growth Factor beta Regulation of p16 Expression in Human Aortic Smooth Muscle Cells. Circulation 132, 1969–1978, doi:10.1161/CIRCULATIONAHA.114.015023 (2015).

