## Supplementary material for "Targeted Sequencing of the 9p21.3 Region Reveals Association with Reduced Disease risks in Ashkenazi Jewish Centenarians": Fig S1-S5; Table S2-S3

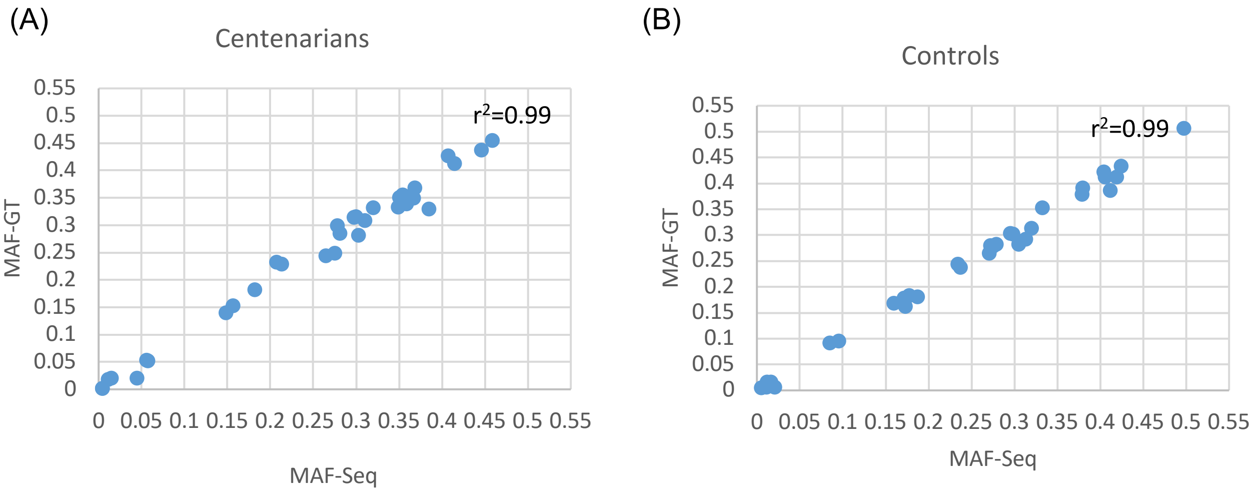


**Figure S1** Genotyping validation of variant allele frequency called from pooled capture sequencing for (A) centenarians and (B) controls. See Table S1 for list of assayed variants.

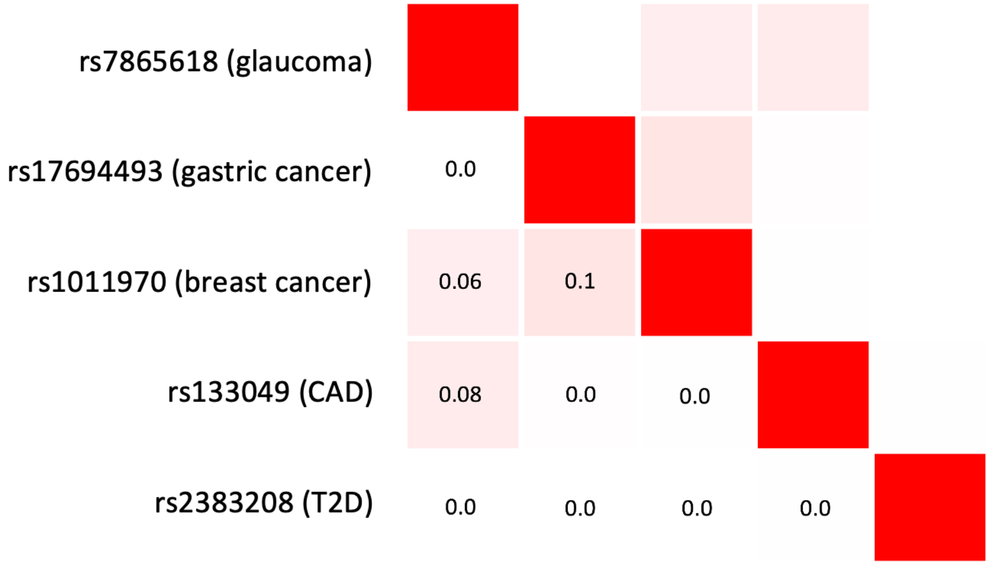


**Figure S2** LD matrix for the five representative GWAS SNPs in all populations from LDLink database. Numbers in the squares indicate R squared correlation.

(A)

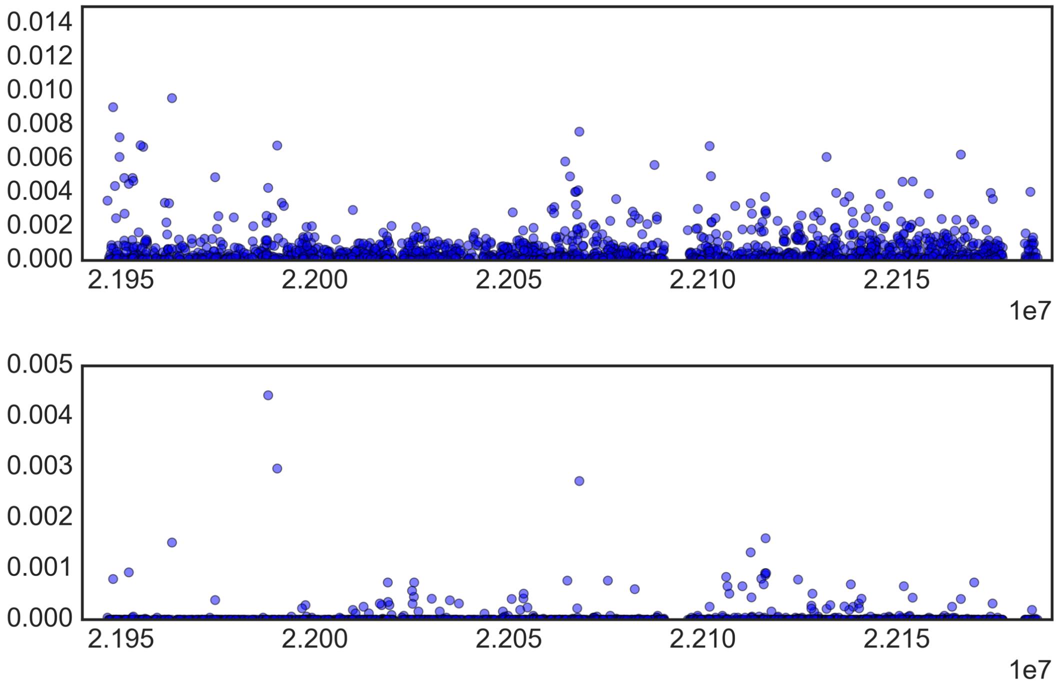

**Figure S3** Feature importance for all variants in (A) random forest classifier and (B) stochastic gradient boosting classifier for centenarian association.

(A)

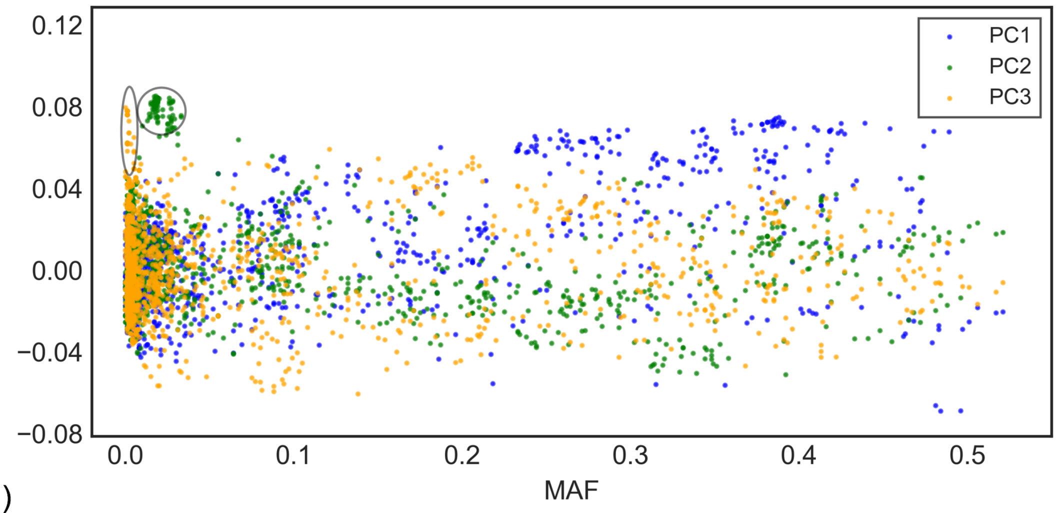


(B)

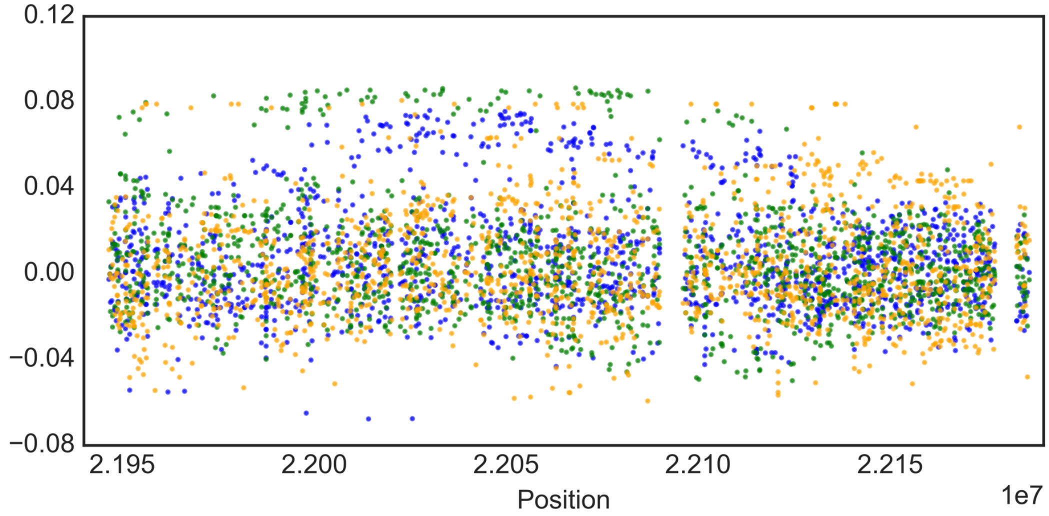


**Figure S4** 2D plots denoting feature weight (y xias) for all SNPs in the first 3 principle components versus their (A) minor allele frequency and (B) position.

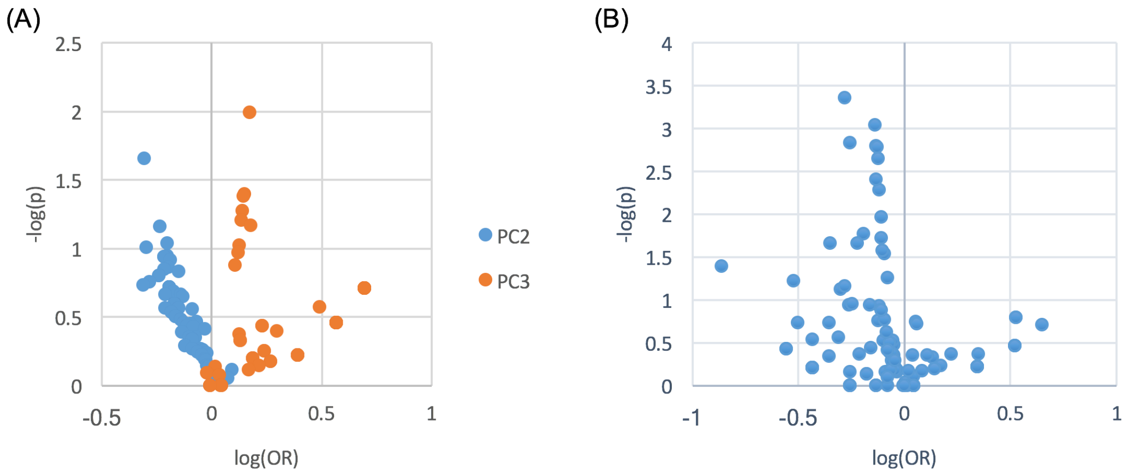


**Figure S5** Volcano plots for (A) important feature SNPs in the principle components 2 and 3 and (B) CDKN2A-downstream SNPs.

**Table S1** List of variants validated by genotyping (attached xlsx separately)

| | MAF-Sequecing | | MAF-Genotyping | |
|---|---|---|---|---|
| ID | Centenarians | Controls | Centenarians | Controls |
| rs111310495 | 0.002 | 0.016 | 0.004 | 0.016 |
| Nov22068072 | 0.002 | 0.016 | 0.004 | 0.012 |
| Nov22089832 | 0.020 | 0.006 | 0.045 | 0.021 |
| rs576966099 | 0.018 | 0.005 | 0.011 | 0.005 |
| rs1011970 | 0.140 | 0.162 | 0.148 | 0.173 |
| rs1063192 | 0.309 | 0.283 | 0.310 | 0.279 |
| rs10738612 | 0.438 | 0.506 | 0.446 | 0.497 |
| rs10757260 | 0.350 | 0.413 | 0.367 | 0.419 |
| rs10757261 | 0.351 | 0.422 | 0.351 | 0.404 |
| rs10757272 | 0.413 | 0.379 | 0.415 | 0.379 |
| rs10811661 | 0.182 | 0.169 | 0.182 | 0.160 |
| rs11515 | 0.153 | 0.183 | 0.157 | 0.178 |
| rs11521166 | 0.053 | 0.092 | 0.056 | 0.085 |
| rs12335941 | 0.339 | 0.413 | 0.359 | 0.406 |
| rs1333039 | 0.299 | 0.244 | 0.278 | 0.235 |
| rs1333040 | 0.332 | 0.302 | 0.320 | 0.299 |
| rs1333042 | 0.369 | 0.353 | 0.369 | 0.333 |
| rs1333049 | 0.456 | 0.434 | 0.459 | 0.424 |
| rs1412829 | 0.281 | 0.265 | 0.303 | 0.270 |
| rs1412832 | 0.229 | 0.181 | 0.213 | 0.187 |
| rs2157719 | 0.314 | 0.280 | 0.298 | 0.272 |
| rs2383207 | 0.356 | 0.313 | 0.355 | 0.320 |
| rs2518722 | 0.244 | 0.304 | 0.265 | 0.295 |
| rs3731197 | 0.330 | 0.386 | 0.385 | 0.412 |
| rs3731211 | 0.249 | 0.282 | 0.275 | 0.305 |
| rs4977574 | 0.427 | 0.392 | 0.407 | 0.379 |
| rs4977756 | 0.286 | 0.238 | 0.281 | 0.237 |
| rs523096 | 0.333 | 0.293 | 0.350 | 0.314 |
| rs72652408 | 0.021 | 0.007 | 0.015 | 0.011 |
| rs74605971 | 0.052 | 0.095 | 0.057 | 0.095 |
| rs75059952 | 0.232 | 0.179 | 0.207 | 0.172 |
| rs7865618 | 0.316 | 0.279 | 0.300 | 0.275 |

**Table S2**: List of all reported GWAS variants in the 9p21.3 sequenced region and their longevity association result.

| | SKAT-O | SKAT | SKAT-C |
|---|---|---|---|
| CDKN2A_downstream | 0.0024* | 0.1380 | 0.0282* |
| CDKN2A | 0.7045 | 0.4285 | 0.1655 |
| CDKN2B | 1.0000 | 0.8812 | 0.4409 |
| CDKN2BAS1 | 1.0000 | 0.8812 | 0.4250 |
| CDKN2BAS1_downstream | 1.0000 | 0.8812 | 0.4212 |
| Total | 0.9904 | 0.8812 | 0.3128 |

**Table S3**: Adjusted p value from SKAT analysis of sequenced region and subdivisions.
